# Post-Stroke Physical Activity Improves Middle Cerebral Artery Dynamic Response

**DOI:** 10.1101/2020.05.13.20100883

**Authors:** Sandra A. Billinger, Alicen A. Whitaker, Allegra Morton, Carolyn S. Kaufman, Sophy J Perdomo, Jaimie L. Ward, Sarah M. Eickmeyer, Stephen X. Bai, Luke Ledbetter, Michael G. Abraham

**Affiliations:** Department of Physical Therapy and Rehabilitation Science, University of Kansas Medical Center, Kansas City, KS; Department of Physical Medicine and Rehabilitation, University of Kansas Medical Center, Kansas City, KS; Department of Neurology, University of Kansas Medical Center, Kansas City, KS; Department of Integrative and Molecular Physiology, University of Kansas Medical Center, Kansas City, KS; Department of Diagnostic Radiology, University of Kansas Medical Center, Kansas City, KS; Department of Interventional Radiology, University of Kansas Medical Center, Kansas City, KS

## Abstract

**Background and Purpose:** The primary aim was to characterize the middle cerebral artery blood velocity (MCAv) dynamic response to an acute bout of exercise at 3- and 6-months post stroke. As a secondary objective, we grouped individuals according to the MCAv dynamic response to the exercise bout as responder or non-responder. We tested whether physical activity, aerobic fitness and exercise mean arterial blood pressure (MAP) differed between groups.

**Methods:** Transcranial Doppler ultrasound measured MCAv during a 90-second baseline (BL) followed by a 6-minute moderate intensity exercise bout. Heart rate (HR), MAP and end tidal CO_2_ (P_ET_CO_2_) were additional variables of interest. The MCAv dynamic response variables included: BL, time delay (TD), amplitude and time constant (τ).

**Results:** Individuals enrolled in the study at 3 months post-stroke and the follow up visit commenced at 6 months post-stroke. Linear mixed model revealed no significant differences in our selected outcomes across between 3- and 6-months post-stroke. Individuals characterized as responders demonstrated a faster TD, higher amplitude, reported higher levels of physical activity and aerobic fitness when compared to the non-responders. No between group differences were identified for BL,τ or exercise MAP. In the non-responders, we observed an immediate rise in MCAv following exercise onset followed by an immediate decline to near BL values while the responders showed an exponential rise until steady state was reached.

**Conclusions:** The MCAv dynamic response profile has the potential to provide valuable information during an acute exercise bout following stroke. Individuals with a greater MCAv response to the exercise stimulus reported regular participation in exercise than those who reported being sedentary.

## Introduction

Impaired cerebrovascular hemodynamic response during exercise has been reported in individuals within one year post-stroke.^1^ We previously demonstrated individuals with middle cerebral artery (MCA) stroke showed no significant differences in resting MCA blood velocity (MCAv) when compared to age- and sex-matched peers.^2^ However, the MCAv dynamic response during exercise was significantly lower for those post-stroke. Despite the impaired cerebrovascular response to exercise, a six-month aerobic exercise treadmill training program in chronic stroke survivors conferred benefit on cerebrovascular reserve for those randomized to the intervention when compared to controls.^3^ This evidence, albeit limited, suggests potential impairment in cerebrovascular control mechanisms following stroke.

The primary objective of this study was to characterize the MCAv dynamic response to a moderate intensity acute exercise bout at 3- and 6-months post stroke. We hypothesized that the dynamic response profile would improve from 3- to 6-months post-stroke. In our previous work comparing young and older adults, we excluded one older female participant from analysis as her data did not fit the exponential model.^4^ Rather than exclude participants with stroke whose data may not fit the exponential model, we decided further scientific inquiry was warranted. As a secondary objective, we divided participants into responders and non-responders based on the model fit. We compared physical activity levels and hemodynamic exercise response. We hypothesized that individuals in the responder group would report greater physical activity, higher estimated *V*□O_2_ maximum and higher mean arterial blood pressure (MAP) during exercise when compared to non-responders.

## Methods

### Participants

We identified individuals post-stroke during the acute or inpatient rehabilitation stay at the University of Kansas Health System. Inclusion criteria were: 1) unilateral ischemic stroke, 2) stenosis in the common and internal carotid artery was <70%.^3,5^ A radiologist (LL) made this interpretation using ultrasound imaging or angiogram in the electronic medical record, 3) 35-95 years of age, 4) physician approval for exercise participation, 5) able to walk > 10 meters without physical assistance and 6) able to travel to the university for the experimental protocol. Exclusion criteria were: 1) unable to consent, 2) inability to perform the exercise movements on the seated recumbent stepper (T5XR NuStep, Inc. Ann Arbor, MI), 3) diagnosis of other neurologic disease, and 4) use of supplemental oxygen.

### Experimental Procedure

Study visits occurred in a laboratory setting at 3- and 6-months post-stroke. Participants were asked to abstain from caffeine for a minimum of six hours, a meal for two hours, and vigorous exercise at least twelve hours prior to the study visit.^4,6^ The University of Kansas Medical Center Human Subjects Committee approved all experimental procedures. Institutionally approved written informed consent was obtained from each individual prior to study participation.

### Familiarization

The laboratory room in which the experimental protocol took place was dimly lit, quiet and maintained a constant temperature between 22-24^°^C.^6^ All external stimuli were kept to a minimum.Participants completed the familiarization session as previously described.^2,4,6^ Following familiarization, we obtained height, weight and completed remaining study documents.

### Protocol Set Up

Individuals were instrumented with the following equipment: 1) transcranial Doppler ultrasound (TCD) (Multigon Industries Inc. Yonkers, NY) and 2-MHz TCD probes for acquisition of MCAv (cm*sec^−1^). The TCD sonographer was blinded to the side of stroke; 2) A 5-lead electrocardiogram (ECG) (Cardiocard, Nasiff Associates, Central Square, NY) recorded HR; 3) beat to beat MAP was obtained from the left middle finger (Finometer, Finapres Medical Systems, Amsterdam, The Netherlands); and 4) nasal cannula and capnograph (BCI Capnocheck Sleep 9004 Smiths Medical, Dublin, OH) assessed end-tidal carbon dioxide (P_ET_CO_2_). We monitored P_ET_CO_2_ to ensure participants did not hyperventilate. Raw data acquisition occurred through an analog-to-digital unit (NI-USB-6212, National Instruments) and custom written software operating in MATLAB (v2014a, The Mathworks Inc. Natick, MA).

### Exercise Protocol

We used moderate intensity exercise (45% to 55% of the participant’s heart rate reserve) for the acute exercise bout. We determined the HR range from one of two equations: 1) age-predicted (220-age) HR maximum (HR_max_) or 2) 164–0.72 × age for participants using beta-blocker medication.^7^

The baseline (BL) recording lasted 90 seconds followed by 6 minutes of moderate intensity exercise. Following the 6-minute exercise bout, the participant engaged in an active cool down for 2 minutes followed by rest until HR, MCAv and MAP returned to BL values. Participants performed a second exercise bout and data points were averaged to optimize signal to noise ratio.^6^ All variables were sampled at 500 Hz and then interpolated to 2.0 Hz. Three-second averages were calculated and then smoothed using a 9-second sliding window average. We used R version 3.2.4 (R Core team, Vienna, Austria)^8^ with the ‘nls’ function package to model the response. To ensure data quality, data with R-to-R intervals greater than 5 Hz or changes in peak blood velocity greater than 10 cm/s in a single cardiac cycle were considered artifact and censored. If acquisitions had more than 15% of data points censored, they were discarded.

### MCAv and Dynamic Response Profile

Kinetics were modeled for MCAv over the entire exercise bout with a mono-exponential model:

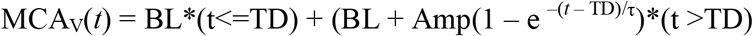

where MCA_V_(*t*) is the MCA_V_ at any point in time, BL is baseline before exercise onset, Amp is the peak amplitude of the response above resting BL, TD is the time delay proceeding the exponential increase in MCA_V_, and is the time constant or time-to-63% of the peak amplitude.

### Six-Minute Walk Test (6MWT)

Participants were given a 20-minute rest following the exercise session. HR and blood pressure were taken to ensure the participant sufficiently recovered from the exercise bout. The 6MWT was performed in a hallway with no distractions or foot traffic according to guidelines outlined by the American Thoracic Society.^9^

### Statistical Analysis

Data are presented as mean ± standard deviation. To account for the missing data across study visits, we used a linear mixed model to test for differences between 3- and 6-months post stroke. To test for between group differences (responders vs non-responders), we performed one-way ANOVA or Mann Whitney U as appropriate following visual inspection of probability plots and Shapiro-Wilk tests. Fisher exact tests were used for categorical data. Differences were considered significant when p < 0.05.

## Results

### Participant Characteristics

We consented 27 participants into the study. One person was consented twice as this individual experienced 2 separate stroke events and has been published as a case report.^10^ Data from the second stroke was not included. Participant demographics (n=26) at enrollment are presented in Table 1. No serious adverse events occurred during any study visit.

**Table 1.** Participant Demographics (n = 26)

|  |  |
| --- | --- |
| Age (years) | 63.0 ± 14.0 |
| Sex % (n) |  |
| Female | 31% (8) |
| Male | 69% (18) |
| Race/Ethnicity* (n) |  |
| White | 18 |
| Black | 8 |
| Native American | 1 |
| Hispanic/Latino | 0 |
| Asian | 1 |
| Pacific Islander | 0 |
| tPA (n) | 14 |
| Thrombectomy (n) | 9 |
| tPA and Thrombectomy (n) | 3 |
| Resting SBP (mm Hg) | 138.5 ± 18.3 |
| Resting DBP (mm Hg) | 80.4 ± 11.2 |
| BMI (kg*(m<sup>2</sup>)<sup>-1</sup>) | 29.8 ± 5.5 |
| Time Post Stroke to 3-month visit (days) | 96.3 ± 12.6 |
| Lesion Size (cm<sup>3</sup>) | 40.2 ± 76.5 |
| Estimated $\dot{V}\dot{O}_2$ maximum (ml*kg<sup>-1</sup>*min<sup>-1</sup>) | 23.2 ± 8.9 |
| Medications (n) |  |
| Beta Blocker | 17 |
| Calcium Channel Blocker | 7 |
| ACE Inhibitor | 5 |
| Angiotensin II Receptor Blocker | 5 |
| Statins | 19 |
| Diuretics | 3 |
| Blood Thinners | 12 |
| Aspirin | 12 |
| Metformin | 5 |
| Insulin | 6 |
| Antidepressants | 9 |

Data are presented as Mean ± SD unless stated otherwise. \*Multiple race categories selected. tPA = Tissue Plasminogen Activator; SBP = Resting Systolic Blood Pressure; DBP = Resting Diastolic Blood Pressure; BMI = Body Mass Index; $\dot{V}\dot{O}_2$ = oxygen uptake; ACE= Angiotensin Converting Enzyme.

Eighteen participants (72.2% male) returned for the 6-month visit. Participants were 65.7 + 14.1 years of age and 188.4 + 14.1 days post-stroke. 83.3% identified as Caucasian, 0% Hispanic,22.2% African American and 5.6% Native American. Multiple race categories were selected. Reasons for loss to follow up included: unable to contact (n = 2); participant’s son had a stroke (n = 1); hospitalized (n = 1); visit scheduled but did not attend study visit (n = 2); no transportation (n = 1) and participant died (n = 1).

### MCAv Dynamic Response Across Time

#### 3-Month Visit

Of the 26 participants, four individuals were excluded from analysis due to: a valid MCAv signal was not acquired, n = 3 and poor MCAv signal acquisition during exercise,n=1. Data are presented for 22 participants.

#### 6-Month Visit

Of the 18 individuals who returned, we excluded 3 datasets from analysis. Reasons for exclusion included an invalid MCAv signal, n = 2 and poor MCAv signal acquisition during exercise, n = 1. Statistical analyses were conducted on 15 datasets.

The MCAv BL and kinetic measures during moderate intensity exercise showed no differences over time for the stroke affected and non-stroke affected MCAv (Table 2). We found no differences between race (p = 0.09) or sex (p = 0.21) for all MCAv data and hemodynamic data (blood pressure, heart rate and P_ET_CO_2_) at rest and during exercise at either 3- or 6-month visits.

**Table 2.**
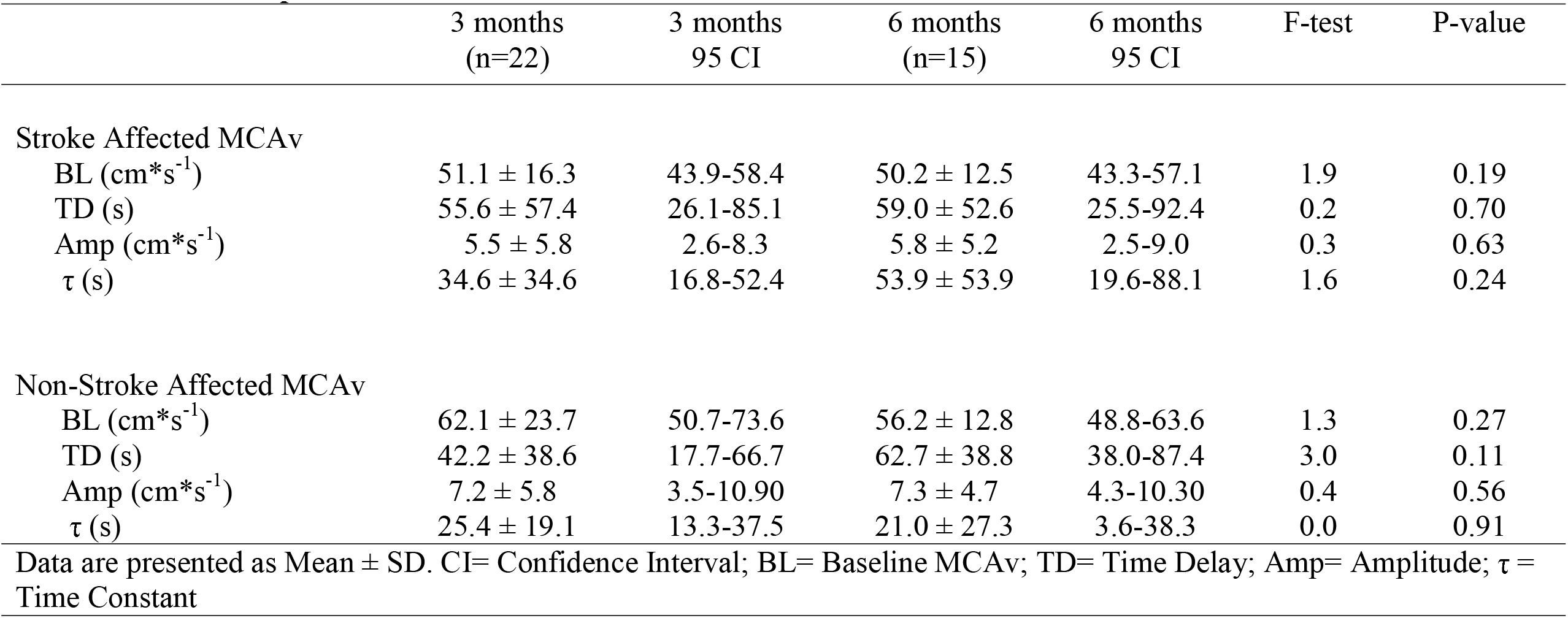
Kinetic Response Across Time

### Responders and Non-Responders (3 Months Post-stroke)

#### MCAv Dynamic Response

Table 3 presents demographics for each group. We observed 12 participants whose MCAv increased in a close-to-exponential pattern being well-fit by a delay + exponential function (responders). The remaining 10 individuals (non-responders) demonstrated poor model fit with visual inspection (n = 5), non-exponential response (n=4) or MCAv was below BL (n=1) after exercise onset on the stroke affected side. On the non-stroke affected side, MCAv decreased below BL after exercise onset for the same individual (n=1) and we observed a non-exponential response in the other participants (n = 9). No between group differences were reported for age, body mass index, 6MWT or the MoCA. The responders were more likely to report taking a statin medication, had greater physical activity levels and had higher estimated *V*□O_2_ maximum values (p <0.05).

**Table 3.**
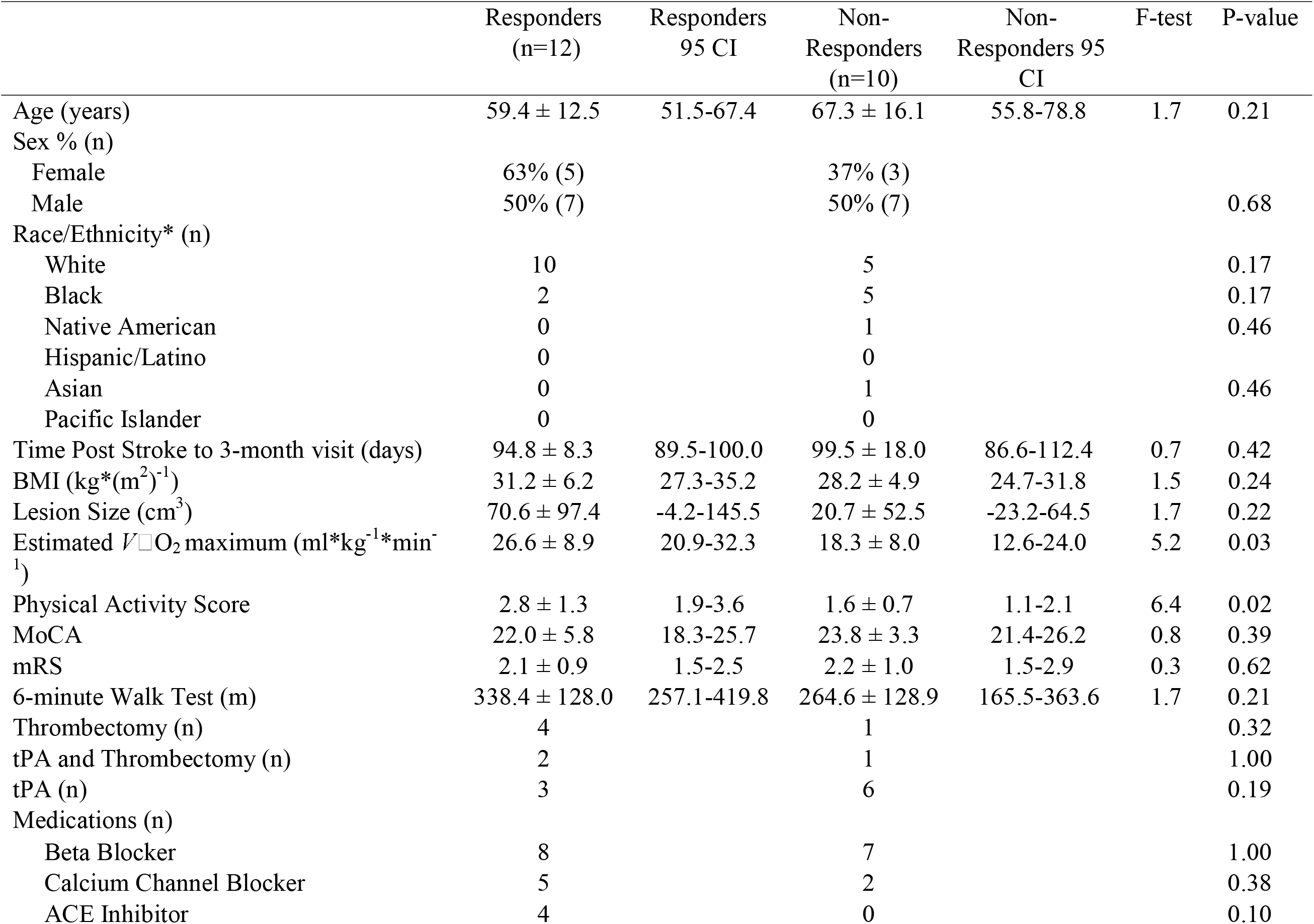
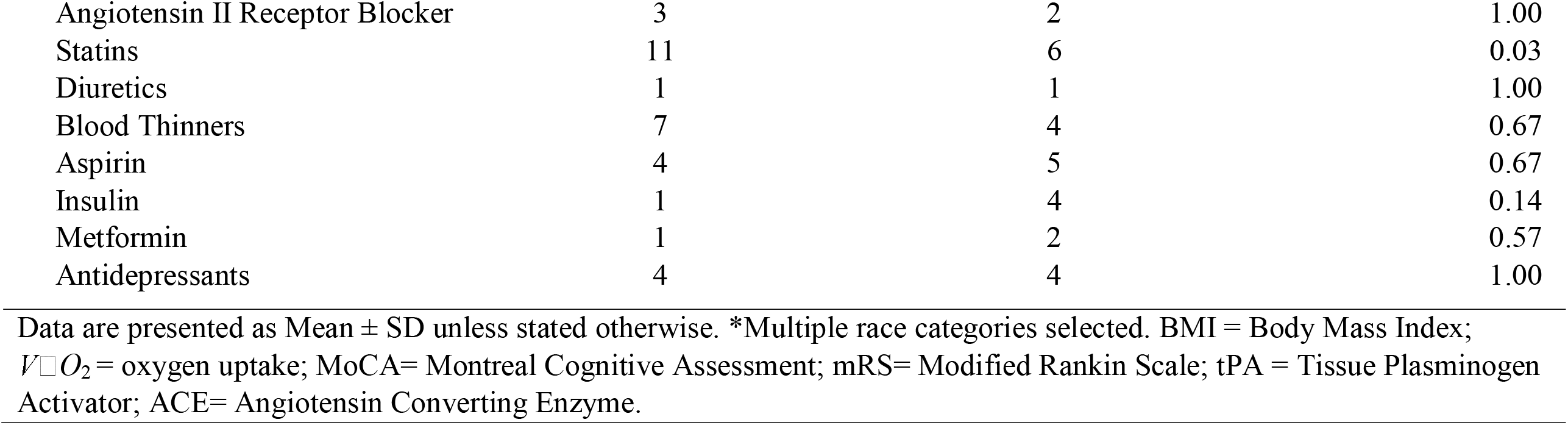
Characteristics of Responders and Non-Responders

Figure 1 shows the BL and MCAv response after the onset of moderate intensity exercise for responders and non-responders. Data from the MCAv kinetics analysis are presented in Table 4. During seated rest prior to exercise onset, BL data for the stroke affected and non-stroke affected MCAv were not significantly different between groups. After exercise onset, MCAv amplitude on the stroke affected side was significantly reduced in the non-responders. This finding is expected based on group assignment (responder/non-responder). For the 5 participants with poor model fit, we observed a significantly slower TD response for the non-responders on the stroke affected side while the τ was not different. Only one person in the non-responder group had data for analysis that fit the model, although a negative amplitude (below BL). The remaining participants (n=9) showed a non-exponential response.

**Table 4.**
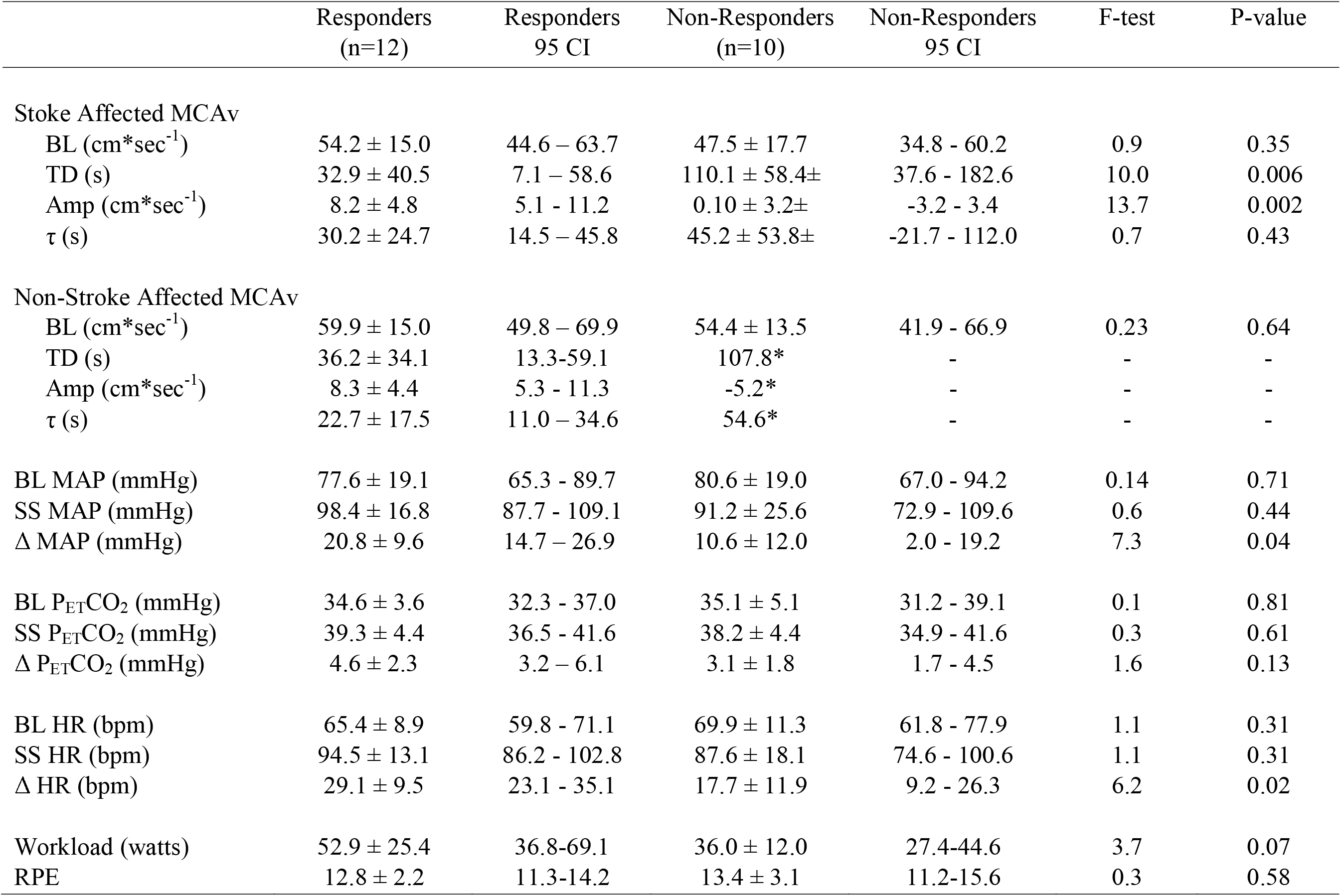
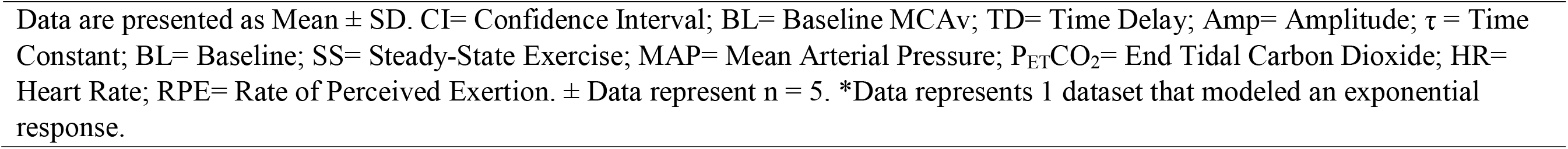
MCAv Kinetics and Exercise Parameters For Responders and Non-Responders

|  | Responders<br>(n=12) | Responders<br>95 CI | Non-Responders<br>(n=10) | Non-Responders<br>95 CI | F-test | P-value |
| --- | --- | --- | --- | --- | --- | --- |
| Stoke Affected MCAv |  |  |  |  |  |  |
| BL (cm*sec<sup>-1</sup>) | 54.2 ± 15.0 | 44.6 – 63.7 | 47.5 ± 17.7 | 34.8 - 60.2 | 0.9 | 0.35 |
| TD (s) | 32.9 ± 40.5 | 7.1 – 58.6 | 110.1 ± 58.4± | 37.6 - 182.6 | 10.0 | 0.006 |
| Amp (cm*sec<sup>-1</sup>) | 8.2 ± 4.8 | 5.1 - 11.2 | 0.10 ± 3.2± | -3.2 - 3.4 | 13.7 | 0.002 |
| τ (s) | 30.2 ± 24.7 | 14.5 – 45.8 | 45.2 ± 53.8± | -21.7 - 112.0 | 0.7 | 0.43 |
| Non-Stroke Affected MCAv |  |  |  |  |  |  |
| BL (cm*sec<sup>-1</sup>) | 59.9 ± 15.0 | 49.8 – 69.9 | 54.4 ± 13.5 | 41.9 - 66.9 | 0.23 | 0.64 |
| TD (s) | 36.2 ± 34.1 | 13.3-59.1 | 107.8* | - | - | - |
| Amp (cm*sec<sup>-1</sup>) | 8.3 ± 4.4 | 5.3 - 11.3 | -5.2* | - | - | - |
| τ (s) | 22.7 ± 17.5 | 11.0 – 34.6 | 54.6* | - | - | - |
| BL MAP (mmHg) | 77.6 ± 19.1 | 65.3 - 89.7 | 80.6 ± 19.0 | 67.0 - 94.2 | 0.14 | 0.71 |
| SS MAP (mmHg) | 98.4 ± 16.8 | 87.7 - 109.1 | 91.2 ± 25.6 | 72.9 - 109.6 | 0.6 | 0.44 |
| Δ MAP (mmHg) | 20.8 ± 9.6 | 14.7 – 26.9 | 10.6 ± 12.0 | 2.0 - 19.2 | 7.3 | 0.04 |
| BL P<sub>ET</sub>CO<sub>2</sub> (mmHg) | 34.6 ± 3.6 | 32.3 - 37.0 | 35.1 ± 5.1 | 31.2 - 39.1 | 0.1 | 0.81 |
| SS P<sub>ET</sub>CO<sub>2</sub> (mmHg) | 39.3 ± 4.4 | 36.5 - 41.6 | 38.2 ± 4.4 | 34.9 - 41.6 | 0.3 | 0.61 |
| Δ P<sub>ET</sub>CO<sub>2</sub> (mmHg) | 4.6 ± 2.3 | 3.2 – 6.1 | 3.1 ± 1.8 | 1.7 - 4.5 | 1.6 | 0.13 |
| BL HR (bpm) | 65.4 ± 8.9 | 59.8 - 71.1 | 69.9 ± 11.3 | 61.8 - 77.9 | 1.1 | 0.31 |
| SS HR (bpm) | 94.5 ± 13.1 | 86.2 - 102.8 | 87.6 ± 18.1 | 74.6 - 100.6 | 1.1 | 0.31 |
| Δ HR (bpm) | 29.1 ± 9.5 | 23.1 - 35.1 | 17.7 ± 11.9 | 9.2 - 26.3 | 6.2 | 0.02 |
| Workload (watts) | 52.9 ± 25.4 | 36.8-69.1 | 36.0 ± 12.0 | 27.4-44.6 | 3.7 | 0.07 |
| RPE | 12.8 ± 2.2 | 11.3-14.2 | 13.4 ± 3.1 | 11.2-15.6 | 0.3 | 0.58 |

Data are presented as Mean ± SD. CI= Confidence Interval; BL= Baseline MCAv; TD= Time Delay; Amp= Amplitude; $\tau$ = Time Constant; BL= Baseline; SS= Steady-State Exercise; MAP= Mean Arterial Pressure; $P_{ET}CO_2$ = End Tidal Carbon Dioxide; HR= Heart Rate; RPE= Rate of Perceived Exertion. ± Data represent n = 5. \*Data represents 1 dataset that modeled an exponential response.

**Figure 1.**
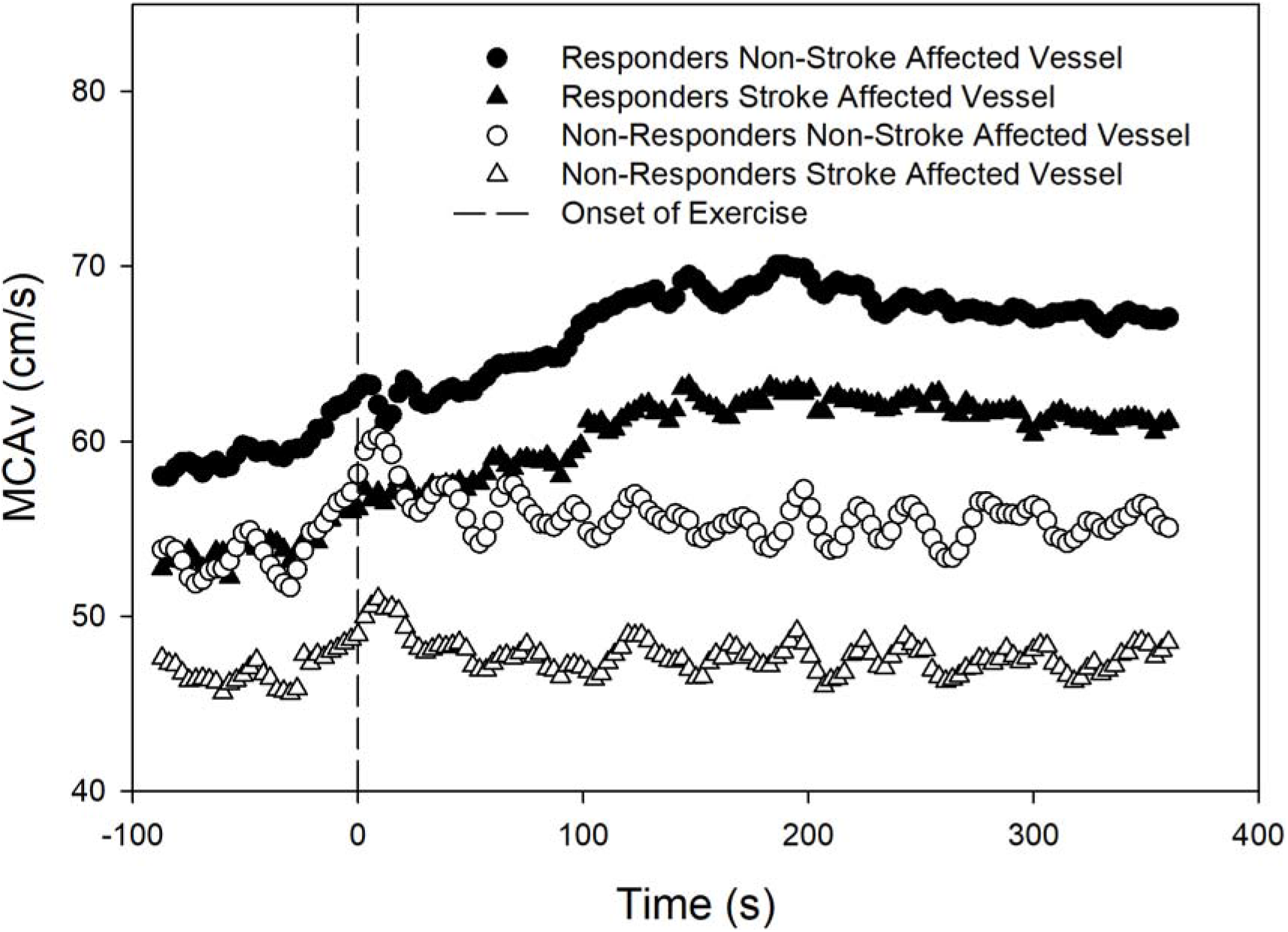
Responders versus Non-Responders, Stoke Affected and Non-Stroke Affected MCAv response to moderate intensity exercise

#### Baseline and Exercise Response in MAP, P_ET_CO_2_ and HR

The baseline and exercise-induced responses (Δ) in MAP, P_ET_CO_2_ and HR are shown in Table 4. The BL and steady state exercise for MAP, P_ET_CO_2_ and HR were not significantly different. We found between group differences for ΔMAP and ΔHR (p < 0.025) but not ΔP_ET_CO_2_ (p = 0.22).

### Responders and Non-Responders (6 Months Post-stroke)

All individuals characterized as a responder at the 3-month visit remained at the 6-month visit. Three individuals initially characterized as a non-responder met the criteria for responder. Of these three individuals, two increased their physical activity score from a 2 (low intensity walking for 10 minutes or less) to a 5 (vigorous activity). The other individual remained in the category of 2 for both visits. Three individuals changed medications related to blood pressure, anticoagulation or antiplatelet therapy that may have influenced MCAv response or other hemodynamic measures at rest or during exercise. In the responder group, one person discontinued clopidogrel. In the non-responder group, lisinopril was added to the medication list while another person reported discontinued use of lisinopril, furosemide and clopidogrel.

There were no group differences between responders (n = 10) and non-responders (n = 5) for age (p = 0.06), time post-stroke (6-month visit, p = 0.47), BMI (p = 0.58), MoCA score (p = 0.54), mRS (p = 0.40) or self-reported physical activity score (p = 0.10). We report significant differences for the non-exercise estimate for *V*□O_2_ maximum (p < 0.01) and six-minute walk test (p = 0.02).

### MCAv Dynamic Response

Responders worked at a higher workload (p = 0.04) with no significant differences in the rate of perceived exertion (p = 0.96). BL MCAv was not different between groups for the stroke affected (p = 0.58) and non-stroke affected MCA (p = 0.87). For the stroke affected MCA, the non-responders showed a significantly slower TD (p = 0.04) and lower amplitude (p = 0.04) while τ was not different (p = 0.93). For the non-stroke affected MCA, no between group significant differences were found for any MCAv dynamic response variables.

### Baseline and Exercise Response in MAP, P_ET_CO_2_ and HR

No between group differences were found for resting and exercise hemodynamic responses, ΔMAP, ΔHR or Δ P_ET_CO_2_.

## Discussion

Aerobic exercise, identical to the intensity used in this study, is recommended for overall health post-stroke.^11^ This study provides novel and relevant information regarding cerebrovascular response to moderate intensity exercise for individuals at 3- and 6-months post-stroke. The main primary findings of the current investigation are: 1) MCAv kinetic response was stable from 3 to 6 months post-stroke for bilateral MCA and 2) MCAv kinetic response (amplitude and TD) significantly differed for those with higher estimated *V*□O_2_ maximum. This finding was consistent for the stroke affected MCA at 3- and 6-months.

### MCAv Dynamic Response Across Time

The primary objective of this study was to characterize the MCAv response at 3- and 6- months post-stroke. To our knowledge, this longitudinal cerebrovascular assessment at rest and in response to exercise had not been previously conducted. We hypothesized that the MCAv dynamic response profile would improve from 3 to 6 months post-stroke. This rationale was developed from published work highlighting improved longitudinal changes in aerobic fitness at 1, 3 and 6 months post-stroke^12^ and aerobic fitness positively influences peripheral vascular health measures in individuals post-stroke.^13^ Our hypothesis that MCAv dynamic response would improve with time was not supported by our findings. We have reported high reproducibility within older adults for resting and exercise MCAv^14^ and bilateral reproducibility for the MCAv kinetic measures.^6^ Therefore, measurement error from our sonographers (SP, CK) was unlikely.

### MCAv Dynamic and Hemodynamic Responses 3 Months Post-Stroke

During exercise onset, neural activation increases the metabolic requirements of the brain above resting values in order to maintain normal function^15–18^ and this demand is met with increased cerebral blood flow. Our laboratory^6,19^ and others^17,18,20^ have shown that moderate intensity continuous exercise produces the greatest increase in MCAv compared to low and high intensity (above anaerobic threshold) continuous exercise.

Ten of the twenty-one individuals enrolled demonstrated a non-exponential MCAv response (non-responders) following exercise onset. This could be indicative of slowed vasomotor control within the cerebrovasculature as both sides showed slower MCAv kinetics or non-exponential MCAv increase.^4^ When considering potential factors, the data in Table 3 show no between group differences for age, sex, race or ethnicity, time of study visit, or the calculated lesion size (SXB) from the electronic medical record.^21,22^ We found responders had greater statin use, self-reported physical activity levels and estimated *V*□O_2_ maximum values. One previous study grouped healthy young adults based on self-reported physical activity levels into low and high fitness.^23^ Those in the high fitness group demonstrated a greater change in MCAv in response to rebreathing-induced hypercapnia while no resting MCAv differences were found. This finding, combined with the present study, underscores the value in considering dynamic challenges such as exercise. Further, others have reported that physically active individuals with higher *V*□O_2_ maximum values exhibit better cerebral hemodynamics than sedentary individuals.^24,25^ Although the evidence reporting the dynamic cerebrovascular response to exercise in stroke is limited,^1,2,6,10^ the present study suggests that physical activity and exercise post-stroke may confer some benefit to overall cerebrovascular health. We acknowledge individuals in the responder group also reported a significantly higher use of statin medications. One study showed 3 months of statin treatment after stroke improved cerebrovascular reactivity compared to the control group while no differences in resting cerebral blood flow were reported.^26^ Our planned future work aims to test whether aerobic exercise interventions can increase the MCAv kinetic response profile and other cerebrovascular outcomes after stroke.

The BL values and steady state hemodynamic responses during the exercise bout were not significantly different between groups. We report between group differences for ΔMAP and ΔHR. We acknowledge that these differences may be largely driven by the significantly higher workload in the responder group, which likely influenced the MCAv amplitude response. One individual (responder) who exercised at 110 watts largely influenced the difference in work rate. He was an avid cyclist prior to his stroke and interestingly, had the largest stroke lesion at admission to the hospital. We report no between group differences for BL, steady state or ΔP_ET_CO_2_. We are confident that the lower MCAv amplitude in the non-responders is not the result of hyperventilation. We closely monitor P_ET_CO_2_ during the exercise bout to ensure our participants stay within the prescribed HR range without hyperventilation.^6^

The literature is well-established that MAP increases with age^27^ and after stroke,^11^ which provides unique opportunities to study exercise-induced changes in MAP and HR. The cerebrovasculature and the associated neuroprotective mechanisms have the ability to autoregulate and buffer the sudden acute increases in MAP associated across various exercise intensities.^28^ As observed in Figure 1, the non-responders show a rapid rise in the MCAv response to exercise followed by an immediate decline with steady state values near BL. One proposed hypothesis for this unexpected response may be an impairment in the cerebrovascular control mechanisms following stroke.^6^ Despite the increased neural activation with exercise and demand for energy, MCAv did not increase. Another plausible explanation may be an impairment in the feed-forward signals coming from the motor cortex to increase MCAv concomitantly with neuromuscular innervation.^4,29^ Our data extend current knowledge regarding cerebrovascular response to exercise and is timely given the increased interest and concern for the role of high-intensity interval training on cerebrovascular response, especially for those with neurological conditions such as stroke.^28,30,31^ The authors of these review papers^28,31^ and a recent study describing the cerebrovascular response to a 30-second bout of high intensity exercise^30^ suggest that caution should be taken when implementing this type of protocol, as the evidence for safety with respect to the brain and its delicate cerebrovasculature has not been tested. A recently published clinical practice guideline for physical therapy suggests higher locomotor intensities benefit walking speed and endurance in chronic stroke.^32^ The current knowledge of cerebrovascular control during an acute exercise bout, especially following stroke, is still in its infancy and warrants further investigation.

### MCAv Dynamic and Hemodynamic Responses 6 Months Post-Stroke

In the chronic phase of stroke recovery, the responders continued to exercise at higher workloads, which may have influenced cerebrovascular response during the exercise bout. At this study visit, only differences in TD and amplitude on the stroke affected vessel remained. All other MCAv kinetic variables and hemodynamic responses were not significant. The reduced sample size warrants caution when interpreting the findings. A noteworthy observation is two of the three newly characterized responders had increased their self-reported physical activity. Whether the increase in physical activity was the primary driver of the higher MCAv amplitude responses remains an important query for future work.

Several limitations in the study design should be considered. First, as with all studies using transcranial Doppler ultrasound, the assumption of constant MCA diameter is important in order for the MCA_V_ to be used as a direct proxy for cerebral blood flow.^6^ Second, maximal exercise testing was not performed to determine maximal heart rate to more accurately prescribe the moderate intensity exercise session. However, as evidenced by the increase in P_ET_CO_2_ during exercise, it is unlikely that individuals exercised above anaerobic levels. Third, we observed an increase in BL MCAv prior to exercise onset. This could be an anticipatory response or nervousness to our verbal instructions 15 seconds prior to exercise onset.

## Conclusion

The cerebrovascular response to acute exercise remained stable between 3- and 6-months post-stroke. Physical activity participation may positively influence the MCAv dynamic response during exercise. Further exploration into the cerebrovascular control mechanisms or other stroke-specific characteristics that may result in a diminished response (non-responders) to exercise and various intensities is needed. The MCAv kinetic response may better inform future exercise trials designed to benefit overall brain health.

## Data Availability

The data are available upon request from the corresponding author.

## Acknowledgments

We would like to thank Yumei Liu, MD, PhD and Emily Nardo for their assistance with data collection and Madison Henry, SPT for her assistance with data organization.

## Sources of funding

Dr. Billinger was supported by the Eunice Kennedy Shriver National Institute of Child Health and Human Development (K01HD067318) and the Wohlgemuth Faculty Scholar Award. Dr. Whitaker and Ms. Morton were supported in part by T32HD057850 from the Eunice Kennedy Shriver National Institute of Child Health and Human Development. Ms. Morton received support from aStudent Scholarship in Cerebrovascular Disease and Stroke (American Heart Association). Dr. Perdomo received partial support from the University of Kansas Alzheimer’s Disease Center (P30AG035982), the KU Medical Center Biomedical Research Training Program and from the National Institute on Aging Diversity Supplement to R01 AG058162. REDCap at University of Kansas Medical Center is supported by CTSA Award (UL1TR000001) from NCRR and NCATS. This work was supported from NCATS awarded to the University of Kansas for Frontiers: Clinical and Translational Science Institute (UL1TR002366) The contents are solely the responsibility of the authors and do not necessarily represent the official views of the NIH or NCATS.

## Disclosures

SAB reports a patent pending (18KU028M-02). MA reports consulting for Stryker Neurovascular and Penumbra Inc. outside the submitted work. AW, AM, CK, SP, JW, SE, SB, and LL report no conflicts of interest. are solely the responsibility of the authors and do not necessarily represent the official views of the NIH or NCATS.

## Notes

### Clinical Trial

This was an observational study and was not registered.

